# Exploring emergency department attendance patterns during the UEFA European Football Championship 2024 in Germany

**DOI:** 10.64898/2026.06.08.26355151

**Authors:** Nesrine Charfeddine, Madlen Schranz, Carmen Schlump, Mirjam Rupprecht, Alexander Ullrich, Michaela Diercke, AKTIN Research Group, Juan Estupinan Mendez

**Author notes:** **Correspondence:** Nesrine Charfeddine.

## Abstract

**Background:** Mass gathering events (MGEs) are associated with several public health challenges and may cause a strain on healthcare services. Literature findings on the impact of MGEs on emergency departments (EDs) are heterogeneous.

**Objectives:** To examine shifts in ED attendance characteristics during a major sporting tournament, namely the UEFA European Football Championship 2024 held in Germany.

**Methods:** We conducted a retrospective observational study using ED data from the Emergency Department Data Registry. We compared baseline ED attendance characteristics between the tournament and the reference period, defined as two weeks before and two weeks after the tournament, and between Germany game days and non-Germany game days. Hourly attendance patterns were analysed for all Germany games using a reference range.

**Results:** We included data from 41 EDs, totalling 253,493 attendances during the study period. A 1.57% increase in attendance was observed during the tournament compared to the reference period, with baseline characteristics remaining similar. The median daily attendance within all EDs was slightly lower on Germany game days (4066) compared to non-Germany game days (4128). Modest changes were observed in the hourly attendance on Germany game days, most notable during the last Germany game where a decrease in attendance below the reference range extended over three hours.

**Conclusions:** The observed shifts in ED attendance were minimal, suggesting that no major changes of public health relevance occurred in ED attendance during the tournament. We highlight the utility of using ED data for monitoring and for enhancing the understanding of the public health risks and challenges associated with MGEs.

## 1. Introduction

Emergency departments (EDs) play a major role in the healthcare system as they usually serve as the first point of contact for patients with acute complaints and provide care in life-threatening situations. They additionally address a multitude of chief complaints of different severity levels, and function as a gateway to specialised healthcare services. Given their pivotal role, routine documentation represents a valuable resource to monitor arising public health threats [1, 2], particularly during specific settings such as mass gathering events (MGEs) [3]. The World Health Organization (WHO) describes MGEs as “events that could strain the public health facilities of the host country” [4]. However, MGEs differ in terms of key characteristics such as event type, attendance demographics and volume, as well as alcohol availability, all of which shape the health risks associated with these gatherings [5].

Major sporting events, as a specific type of MGEs, attract large stadium crowds and extensive media attention. Hosting tournaments of this magnitude poses multiple public health challenges. Indeed, large high-density crowds, prolonged close contact between attendees, and surges of national and international travellers constitute a favourable environment for the transmission of communicable diseases (CDs) [6, 7]. Additionally, elements such as the availability and advertisement of alcohol in stadiums and fan zones, extreme weather conditions, physical exertion, and heightened emotional stress during sports tournaments may lead to an increase in the occurrence of non-communicable diseases (NCDs), potentially heightening the demand on EDs [8-10]. These changes have been reported to occur mostly on game days involving the host country as they draw large audiences and spectator engagement [9, 11, 12]. However, international studies report heterogeneous findings regarding the impact of sporting events on emergency healthcare services; while some indicate an additional strain during the tournament [13], others observed no significant change in ED patient volume [12], with reports of a decrease in attendance, particularly on game days [14].

Studies examining changes in ED patient volume and baseline characteristics during major sporting events remain limited in Germany, as they typically focused on specific health indicators rather than overall attendance, leaving a gap in understanding how MGEs may impact ED utilisation. In 2024, the 17^th^ edition of the men’s UEFA European Football Championship (EURO 2024) took place in Germany and welcomed 2.7 million spectators across ten stadiums and 6.2 million in official fan zones [15]. Given the scale of the event, it provided a unique opportunity to examine potential nationwide changes in ED attendances, using routine data from multiple EDs included in the Robert Koch Institute’s (RKI) ED surveillance system.

We aimed in this study to examine changes in ED attendance and characteristics during the EURO 2024 in Germany, with a particular focus on hourly attendance patterns on game days involving the host country. Ultimately, we aim to contribute to the existing body of knowledge on potential public health risks associated with hosting events of this magnitude, thereby supporting improved planning and resource allocation.

## 2. Methods

### 2.1 Study design and data source

We conducted a retrospective observational study to describe ED attendance and baseline characteristics during the EURO 2024. We used routinely collected case-based, anonymized data from the Emergency Department Data Registry (data access request ID 2019-003, 3^rd^ amendment) [16]. Data is transmitted daily to the RKI where it is processed and stored in an internal database. We used the following variables for analysis: gender, age group, ED visited, date and time of visit, disposition and triage levels. Only visits with complete data for gender, age, date and time of visit, as well as the ED visited are transferred to the RKI. Time of visit is categorized by hour such as hour 18:00 represents the interval 18:00 to 18:59. The variable disposition represents the outcome of the patient after ED visit, while triage represents the urgency level.

### 2.2. Study period and study population

The study period spanned from the 24^th^ of May to the 28^th^ of July 2024, encompassing the EURO 2024 (14^th^ of June to 14^th^ of July 2024) and a reference period, each totalling 31 days. The latter consists of a pre-EURO 2024 interval covering 17 days before the tournament (24^th^ of May to 9^th^ of June 2024), and a post-EURO 2024 interval covering 14 days after (15^th^ to 28^th^ of July 2024) **(Supplementary Figure S1)**. This reference period was selected in the same year as the tournament instead of a corresponding period in the previous years to include a maximum number of EDs, given that the number of participating ED in the registry increased over time. This approach additionally allowed to ensure comparability between the periods as it is adjacent to the EURO 2024.

Furthermore, the reference period was aligned with the EURO 2024 in terms of duration and day-of-week distribution as ED attendance and characteristics vary across weekdays [17]. To ensure a similar distribution of weekdays, a four-day gap was introduced between the pre-tournament interval and the tournament. This gap additionally allowed to account for changes occurring in ED attendances due to the surges of travellers prior to the tournament’s start.

We restricted our analyses to EDs that consistently provided daily data for both the tournament and the reference period. We excluded four EDs which exhibited unusually low overall daily visit counts, which were attributed to incomplete data transmission. Ultimately, a total of 41 EDs were included, four of which were paediatric EDs, providing coverage for ten federal states in total: Baden-Württemberg, Bavaria, Berlin, Lower Saxony, North Rhine-Westphalia, Rhineland-Palatinate, Saxony, Saxony-Anhalt, Schleswig-Holstein, and Thuringia. Four of the covered federal states hosted Germany games. All attendances recorded during the study period were included in the analysis.

### 2.3. EURO 2024: tournament structure and game periods definition

The EURO 2024 was held across seven federal states and comprised of 51 games, five featuring Germany’s national team: three in the group stage (14^th^, 19^th^ and 23^rd^ of June 2024), one in a round of 16 (29^th^ of June 2024), and one quarter-final (5^th^ of July 2024). In accordance with previous studies, we focused our analysis on games involving the host country [11, 12]. We defined “Germany game days” as days where the German national team played, and “non-Germany game days” as days where no games were held or games in which Germany did not participate. For each game day, we used previously defined and reported game period definitions: a pre-game period (four hours before game kick-off), a game period representing the duration of the game, and a post-game period (four hours after the end of the game) [12].

### 2.4. Statistical Analysis

First, we conducted descriptive analyses by calculating the overall attendance during the EURO 2024 and the reference period, and by comparing the distributions of age groups, gender, triage and disposition levels. A similar approach was applied for describing attendance throughout the tournament, on Germany game days and non-Germany game days. We reported for each period the median of daily attendance across the included EDs, as well as the interquartile range (IQR) using the 25^th^ and 75^th^ percentiles.

Second, we compared the hourly ED attendance on Germany game days to reference days. We defined “reference days” as days within the reference period matched to a game day by day-of-week. As an example, a game held on a Friday was compared to all Fridays of the reference period. This analysis was conducted in two steps:

The first step consisted of calculating the difference between the hourly attendance on Germany game days and the corresponding hourly mean of reference days. We defined a “relative-to-game hour” as a temporal marker, where hour 0 corresponded to game kick-off, negative hours indicated hours before the game, and positive hours represented hours after the game.

The second step consisted of constructing time series of the hourly attendance on Germany game days. We defined a reference range with upper and lower limits set at two standard deviations above and below the mean of reference days. This analysis was conducted for the overall hourly attendance, as well as stratified by gender and age group.

All analyses and data visualisations were performed using the programming language R (version 4.4.1).

## 3. Results

### 3.1. Study population

We firstly compared the number of attendances and their characteristics between the EURO 2024 and the reference period. A total of 253,493 attendances were analysed, of which 127,732 attendances were recorded during the tournament compared to 125,761 attendances in the reference period, representing 1,971 (+1.57%) additional attendances. The median daily attendance during the EURO 2024 period was slightly higher than the reference period, reaching respectively 4119 (IQR: 3971 - 4260) and 4026 (IQR: 3868 – 4212). Similar proportions of male and female attendances were observed across both periods, indicating no difference in gender distribution, with male attendances accounting for approximatively 52% in both periods. The distribution of age groups was additionally similar, with the highest share of attendances observed among individuals aged 20-39 years and the lowest among those aged 80 years and older. Similarly, the proportions of triage and disposition levels were highly comparable **(Table 1)**. These findings are consistent when stratifying by gender; the distribution of triage and disposition levels did not differ in male and female attendances during the EURO 2024 and the reference period **(Supplementary Figure S2-S3)**.

**Table 1.**
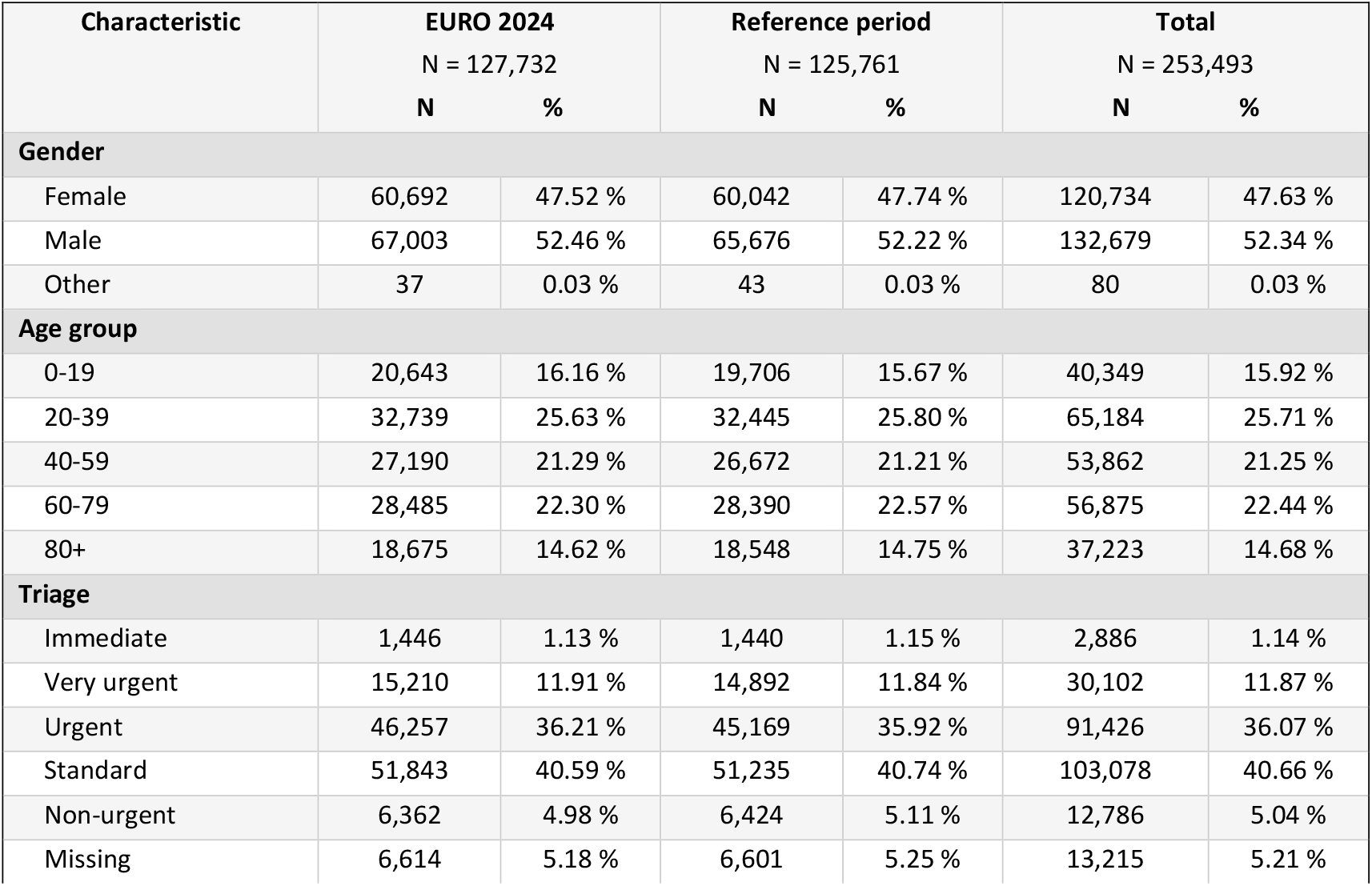
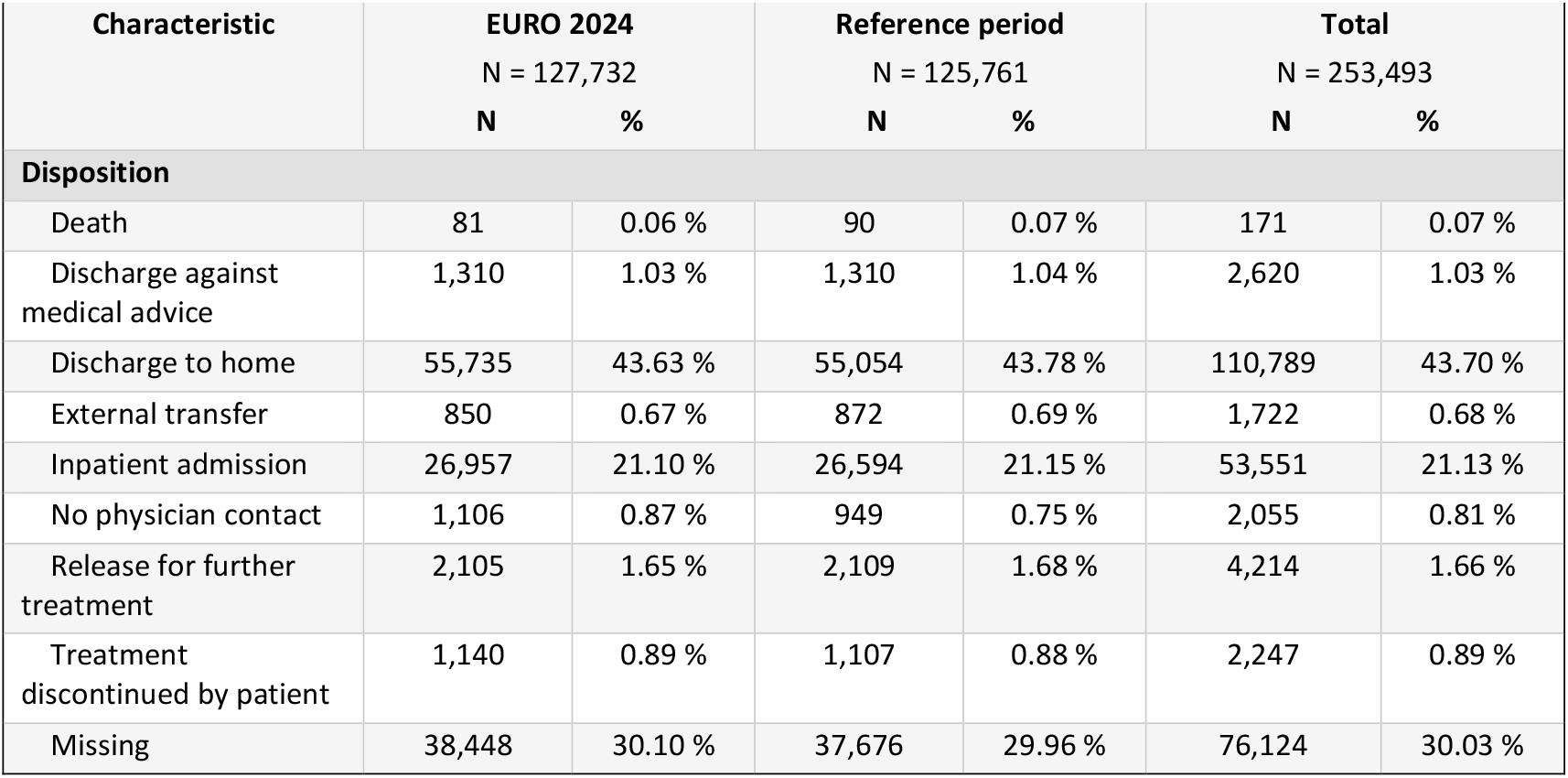
Baseline characteristics of the overall attendance in the 41 included emergency departments during the EURO 2024 and the reference period.

We secondly described attendance characteristics throughout the tournament, comparing Germany game days and non-Germany game days. A total of 20,118 attendances were recorded on the five Germany game days, and 107,614 attendances on non-Germany game days which lasted 26 days. Non-Germany game days included nine days where no games were held, and 17 days where games not involving the host country took place.

The median daily attendance was slightly lower on Germany game days compared to non-Germany game days, reaching respectively 4066 (IQR: 4063 - 4119) and 4128 (IQR: 3958 - 4288). Gender and age group distributions were also comparable across both periods, with male visits and individuals aged 20-39 years being the most represented. The age group 0-19 exhibited the highest difference between the two periods with a 1.51 percentage-point increase in its proportion on Germany game days compared to non-Germany game days, while the most notable decrease was noted among individuals aged 60-79, reaching a 1.20 percentage-point decrease. Additionally, minor differences were observed in the triage and disposition levels. The proportion of the triage level “Urgent” decreased by 0.69 percentage-point on Germany game days. For disposition, the proportion of “Discharge to home” increased by 1.16 percentage-point, while the proportion of “Inpatient admission” decreased by 1.38 percentage-point compared to non-Germany game days **(Table 2)**.

**Table 2.**
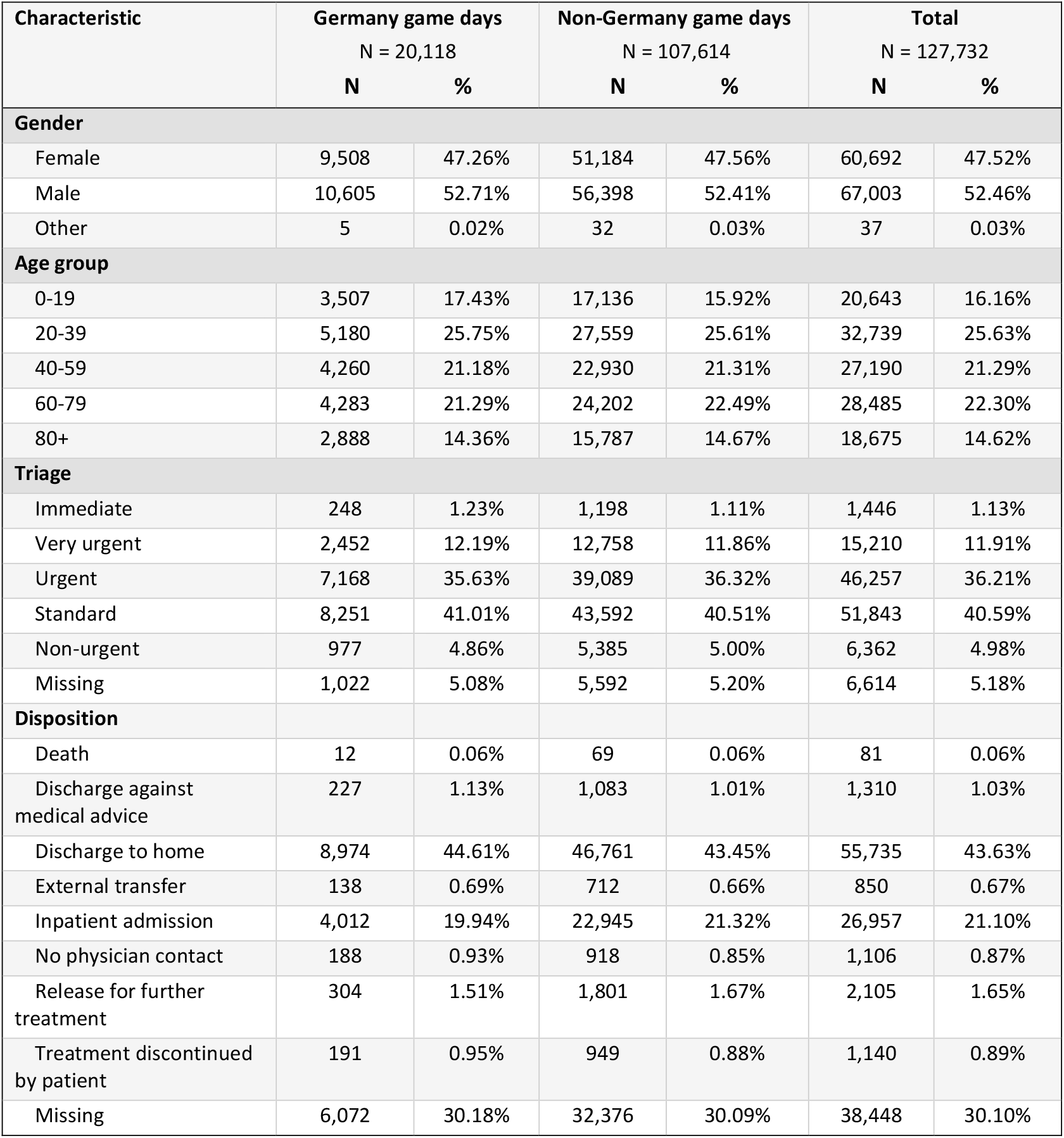
Baseline characteristics of the overall attendances in the 41 included emergency departments on Germany game days and non-Germany game days during the EURO 2024. Germany game days were held across 5 days. Non-Germany game days were held across 26 days.

### 3.2. Hourly attendance analysis

We computed the difference between hourly attendance on Germany game days and the hourly mean of reference days. In the pre-game period, attendances tended to decrease compared to reference days. This decreasing tendency was observed on all Germany game days, with varying magnitude, and span over time. The game held on the 23^rd^ of June additionally exhibited a slight increase in attendance at 4 hours before the game. Game durations varied between approximatively two hours for the first four games and three hours for the last. During the game period, differences in hourly attendance between game days and reference days were heterogenous. On one hand, we noted a decline in attendance on the 14^th^ of June, representing the opening game, on the 19^th^ of June, and on the last Germany game held on 5^th^ of July. On the other hand, modest increases in attendance were observed on the 23^rd^ and 29^th^ of June. The post-game period exhibited an increase in attendance for the games held on the 19^th^ and 29^th^ of June, while the remaining games saw minimal changes ranging from a decrease to a less notable increase **(Figure 1)**.

**Figure 1.**
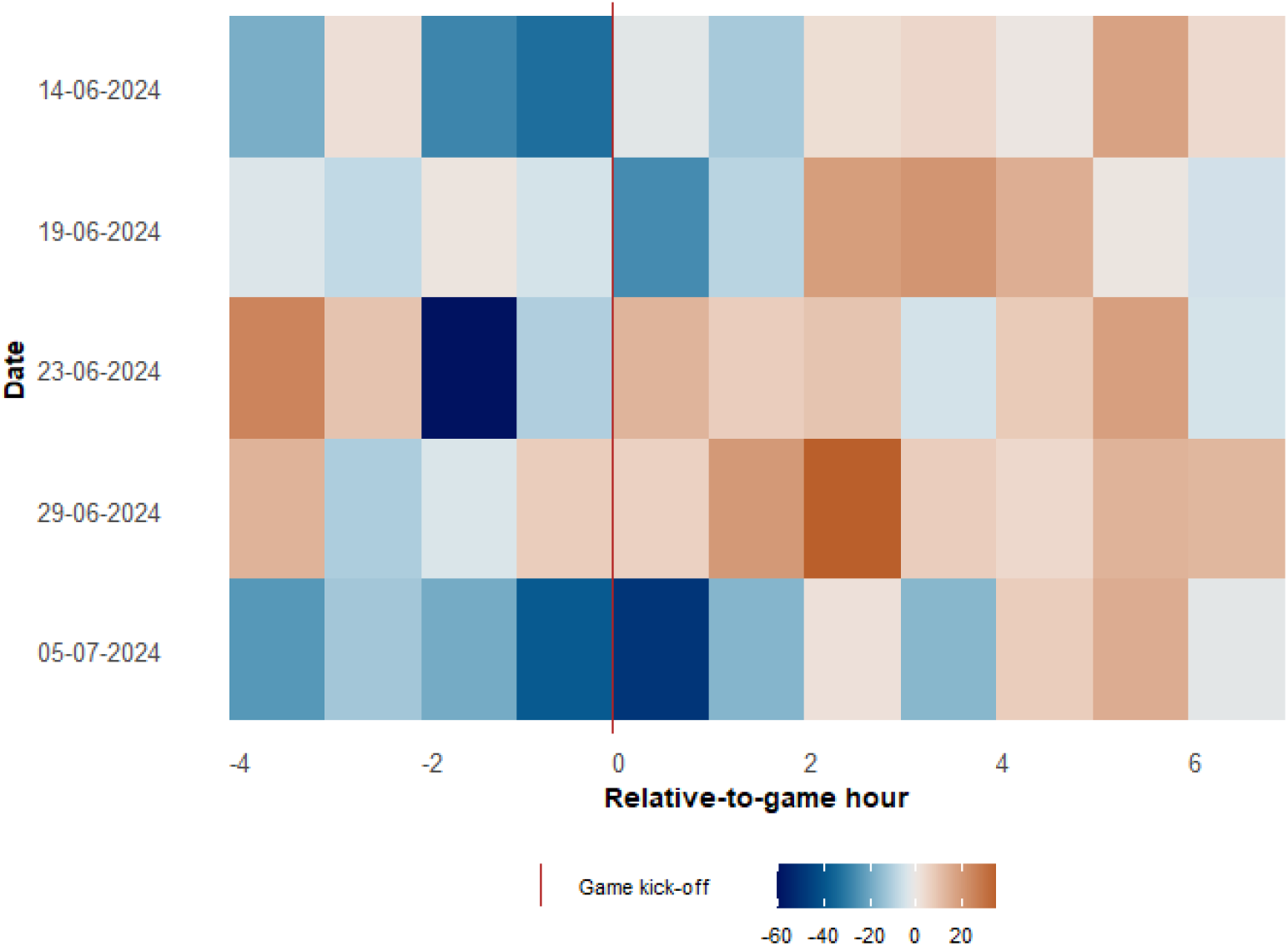
Heatmap representing the differences between the hourly attendance on Germany game days and the hourly mean of reference days. The colour blue indicates a negative difference; orange indicates a positive difference. Colour shades mirror the magnitude of the difference. The X-axis represents relative-to-game hour; 0 marks game kick-off. The game period lasted two hours for the first four Germany games, three for the last game.

We furthermore analysed the hourly attendance patterns using a reference range. We observed the most sustained decrease in attendance on the last Germany game which took place on the 5^th^ of July **(Figure 2)**. This change extended over three hours, decreasing by 5.43 attendances from the reference range at two hours before game kick-off, by 16.57 one hour before game kick-off, and by 15.85 during the first hour of the game. No changes beyond the range occurred in the post-game period. The most pronounced decrease in attendance was noted on the 23^rd^ of June, two hours before the game, reaching a decrease by 34.42 attendances **(Supplementary Figure S4)**. Furthermore, the highest increase in attendance was observed on the 29^th^ of June in the game and post-game periods **(Supplementary Figure S5)**. This increase extended over two hours, reaching an additional 5.21 attendances from the reference range one hour after game kick-off, and 23.34 two hours after game kick-off. The games held on the 14^th^ and 19^th^ of June exhibited less notable shifts.

**Figure 2.**
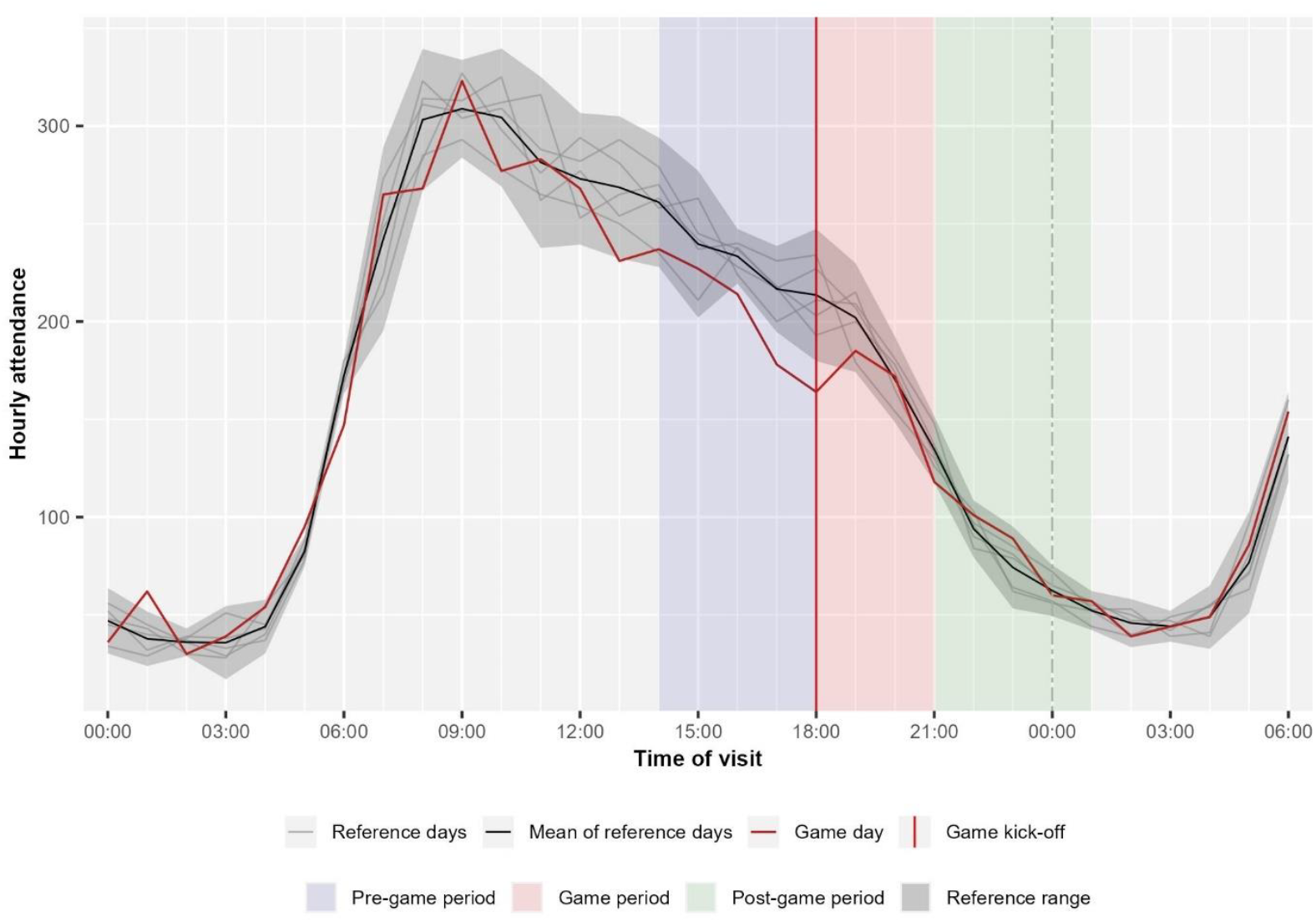
Hourly attendance on the last Germany game held on the 5^th^ of July, compared to a reference range representing two standard deviations above and below the hourly mean of reference days. Game periods are highlighted.

The reported changes in the nationwide hourly attendance pattern were mainly observed in the age-groups 20-39, 40-59 and 60-79 years. However, these changes were minimal and heterogenous across games **(Supplementary Figure S6)**. The age group 20-39 additionally exhibited a slight upward trend in the post-game period beyond the reference range during the games held on the 14^th^ and 19^th^ of June. Furthermore, we observed shifts in the hourly attendance pattern among male and female visitors with varying magnitude during the pre-game and game period **(Supplementary Figure S7)**, while the post-game rise in attendance was more pronounced in male attendances. Indeed, on the game held on the 29^th^ of June, an increase by 17.86 in male attendances two hours after game kick-off was noted, as well as by 6.63 five hours after kick-off, compared to the reference range **(Figure 3)**.

**Figure 3:**
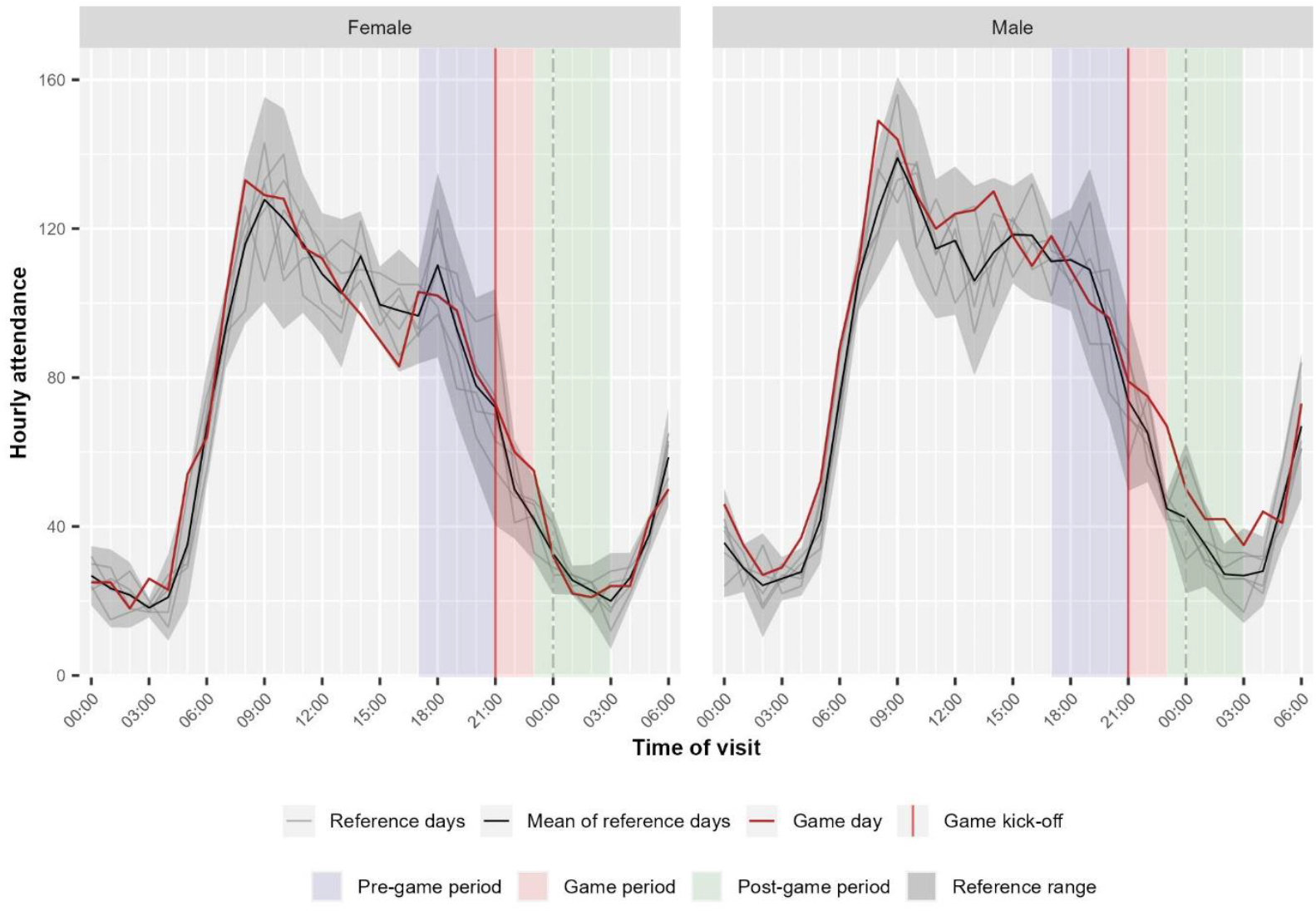
Hourly attendance on the fourth Germany game held on the 29^th^ of June 2024 stratified by gender, compared to a reference range representing two standard deviations above and below the hourly mean of reference days. Game periods are highlighted.

## 4. Discussion

We aimed throughout this study to describe how ED attendance characteristics change during the EURO 2024 compared to a reference period, as well as on Germany game days compared to non-Germany game days, and to analyse shifts in hourly attendance around Germany game kick-offs. Our findings suggest that no major changes in ED attendance occurred during the EURO 2024. We noted a small 1.57% increase in attendance during the tournament compared to the reference period, with similar baseline characteristics. Previous studies documented a more substantial increase in ED patient volume during tournaments, reaching an additional 8% during the 2011 Rugby World Cup [18]. These spikes in ED attendance have often been attributed to an increase in NCD-related attendances, such as in alcohol intoxication and myocardial infarction [9, 10, 18].

Furthermore, we report that throughout the EURO 2024, the median daily attendance was slightly lower on Germany game days compared to non-Germany game days, reaching respectively 4066 (IQR: 4063 - 4119) and 4128 (IQR: 3958 - 4288). Although modest, our findings align with some reports in the literature which suggest no additional strain on EDs during sporting events, with some reports describing a significant decrease in attendance on game days [19, 20]. These changes have been reported particularly during regional and international tournaments, primarily among male attendances [21]. Conversely, we did not observe a change in the proportion of male attendances between Germany game days and non-Germany game days. Nonetheless, findings in the literature are heterogeneous, with reports of more substantial increases in ED attendance on game days [22].

The influence of sporting events on EDs is, however, not limited to changes in the number of visits. It can also be reflected by shifts in patients’ clinical characteristics. McGreevy et al. reported that, while no correlation was found between sporting events and total ED attendances, a significant increase in hospital admissions was observed [20]. These changes in patients’ characteristics can pose additional strain on EDs, as complex and severe cases require more extensive and prolonged care. Conversely, in our study, the distribution of triage and disposition levels of ED attendances on the EURO 2024 and the reference period were similar. Prominently, Germany game days and non-Germany game days exhibited minor differences; an increase by 1.16 percentage-point in the proportion of patients discharged to home was noted, as well as a decrease by 1.38 percentage-point in the proportion of inpatient admission, suggesting that a slightly higher proportion of milder cases was observed on game days involving the host country.

We hypothesise that the availability of on-site medical care may have contributed to the absence of anticipated increase in ED attendance during the EURO 2024 and on Germany game days. The German Red Cross reported a total of 4,768 interventions in stadiums and fan zones during the EURO 2024, the majority of which were attributed to cardiovascular conditions [23]. Although our study does not directly assess the effect of on-site medical care, the large number of interventions conducted on-site highlights its importance for the safe conduct of tournaments, as it may lower the strain on healthcare services by enabling rapid response, treatment of milder cases and transport to appropriate healthcare facilities [24].

To further investigate ED attendance changes during the tournament, we examined differences in the hourly attendance patterns between Germany game days and the reference period. Although minimal and with varying magnitude, our findings suggest that hourly attendance changed around game kick-off, in line with previous reports that documented changes in the healthcare-seeking behaviour of ED visitors around games. Reported modifications manifested mainly by a decrease in attendance before and during the game, and an increase after the end of the game [12, 14]. It is plausible that the decrease in attendances before and during the event is partly attributable to a decrease in low acuity attendances, as patients may postpone seeking care for mild symptoms before and during games of interest. The spike in attendance following the game may reflect a rebound in attendances, which has been attributed in the literature to an increase in NCD-related attendances, as the emotional and physical stress experienced during games can contribute to an increase in cardiovascular events [25]. The temporal relationship between emotional triggers and the occurrence of these events has been further investigated by Wilbert-Lampen et al. who noted that the highest incidence of cardiovascular events was observed during the first two hours after game kick-off and maintained for several hours [9].

Although not observed on all Germany game days, our analysis revealed a general trend of decreased attendance during the pre-game and game periods and an increase afterwards. Similar trends were reported during the 2016 edition of the EURO, where, despite no changes in ED daily attendance, the hourly attendance patterns shifted on game days [12]. Additionally, in our study, the most notable and sustained change was observed in the last Germany game, which concluded in a loss and elimination from the tournament; this quarter-final recorded the largest audience of the EURO 2024, reaching a total of 29.5 million viewers [15]. Previous findings suggest that the magnitude of changes observed in the hourly attendance patterns varies depending on game intensity and importance; emotionally-charged games such as games in later stages of a tournament and games involving shoot-outs appear to have higher impact on NCD-related visits [11, 26]. These game characteristics may play a higher impact on the occurrence of NCDs than the final outcome of the game, as increases in attendance have been reported following both wins and losses of the host team [9, 27, 28].

While our findings provide insight into how ED attendance characteristics change in light of a major sports tournament, several limitations should be addressed. First, competing events such as school holidays, bank holidays, and other simultaneous MGEs occurring during the study period were not accounted for. Second, the selection of the reference period has inherent limitation; although adjacent to the EURO 2024 and similar in duration and number of weekdays, underlying differences may still be present as the periods were not selected in the same time frame. Third, potential selection bias may arise from the selection of EDs, which was based on data availability and consistency. A total of 41 EDs were included nationwide, representing a relatively low coverage of EDs in Germany which has over 1,000 EDs; however, this study demonstrated that meaningful patterns can still be observed, and analyses will become more robust as coverage expands in the future. Furthermore, the included EDs may not be representative as their participation in the study is voluntary. Notably, analyses restricted to EDs in proximity to the tournament venues were not feasible with the available data. Nevertheless, since public-viewing areas were organised throughout Germany and as major sporting events draw widespread attention, changes in ED demand can be observed not only in EDs near stadiums but also nationwide. Lastly, in line with previous published studies, our analysis investigated changes in the hourly attendance pattern on games involving the host country. However, major sports tournaments such as the EURO 2024 attract international spectators; a change in ED demand may therefore also be observed during non-Germany game days. Notably, games that do not directly involve the host country but influence its progression in the tournament, as well as decisive games in the tournament’s outcome can draw similar spectator engagement and audiences and potentially have an impact on ED demand.

## 5. Conclusion

To the best of our knowledge, a descriptive analysis of overall ED attendance and baseline characteristics during major sporting events in Germany has not been previously published. We report that, first, no major changes in ED attendance patterns and characteristics occurred during the EURO 2024. Second, we noted modest shifts in the hourly attendance pattern around game kick-off, most notably on the last Germany game day. The observed changes were minimal, suggesting that no major changes of public health relevance occurred in ED attendances during the EURO 2024. We highlight the utility of using ED data for monitoring and for enhancing the understanding of the public health risks and challenges associated with hosting MGEs and should be employed as real-time surveillance in future events to provide timely reassurance and inform preventive measures.

## Statements

### Data availability

Aggregated data can be made available upon request after agreement with the Emergency Department Data registry.

### Collaborators

NC, MS and JEM conceptualized the study. NC further developed the study design, conducted the data analysis and created the tables and figures. MS, JEM and AU provided guidance on the methodology. NC wrote the original draft. MS, JEM, CS, AU, MR and MD contributed to editing and revising the manuscript. All authors read and approved the final version.

### Ethical statement

The Emergency Department Data Registry received medical ethical approval from the Ethics Committee of the Otto von Guericke University Magdeburg, Medical Faculty (160/15). For this study, data usage approval was obtained from the AKTIN Data Use and Access Committee (Data access request ID 2019-003, 3^rd^amendment).

### Use of artificial intelligence tools

Grammarly was used to improve the clarity and readability of the manuscript. The OpenAI tool ChatGPT was used to resolve R coding errors and to assist in creating functions for generating figures. All content was reviewed and approved by the authors.

## Acknowledgements

We want to express our gratitude to the AKTIN Network, the Emergency Department Data Registry and all participating emergency departments for providing data for this analysis and for their valuable contributions and feedback for this work. The participating emergency departments connected to the Emergency Department Data Registry are listed on the following website: https://aktin.org/wp-content/uploads/Anlage_1_Studienzentren_2026-04-15.pdf.

