## Supplementary Material for "Exploring emergency department attendance patterns during the UEFA European Football Championship 2024 in Germany"

**Supplementary Figure S1:** *EURO 2024 study period: pre-EURO 2024, EURO 2024, post-EURO 2024 periods definition.*

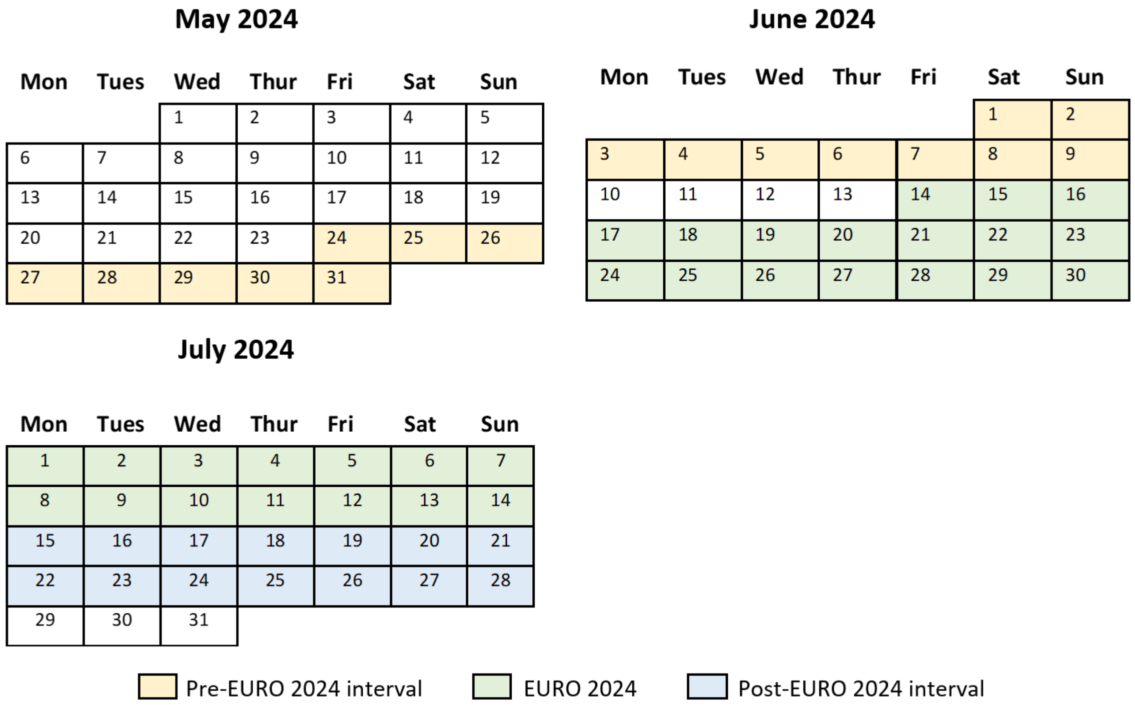

**Supplementary Figure S2:** *Proportion of daily mean attendances by disposition level during the EURO 2024 and the reference period, stratified by gender. Error bars represent two standard deviations above and below the mean.*

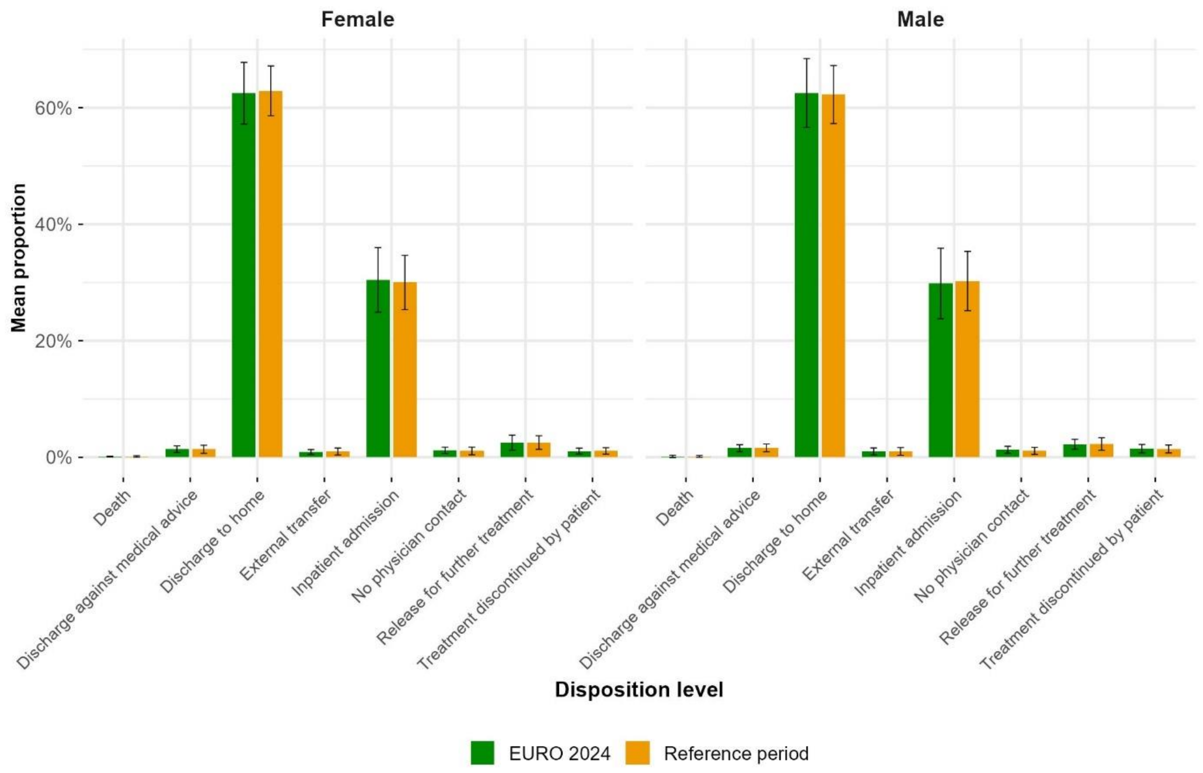


**Supplementary Figure S3:** *Proportion of daily mean attendances by triage level during the EURO 2024 and the reference period, stratified by gender. Error bars represent two standard deviations above and below the mean.*

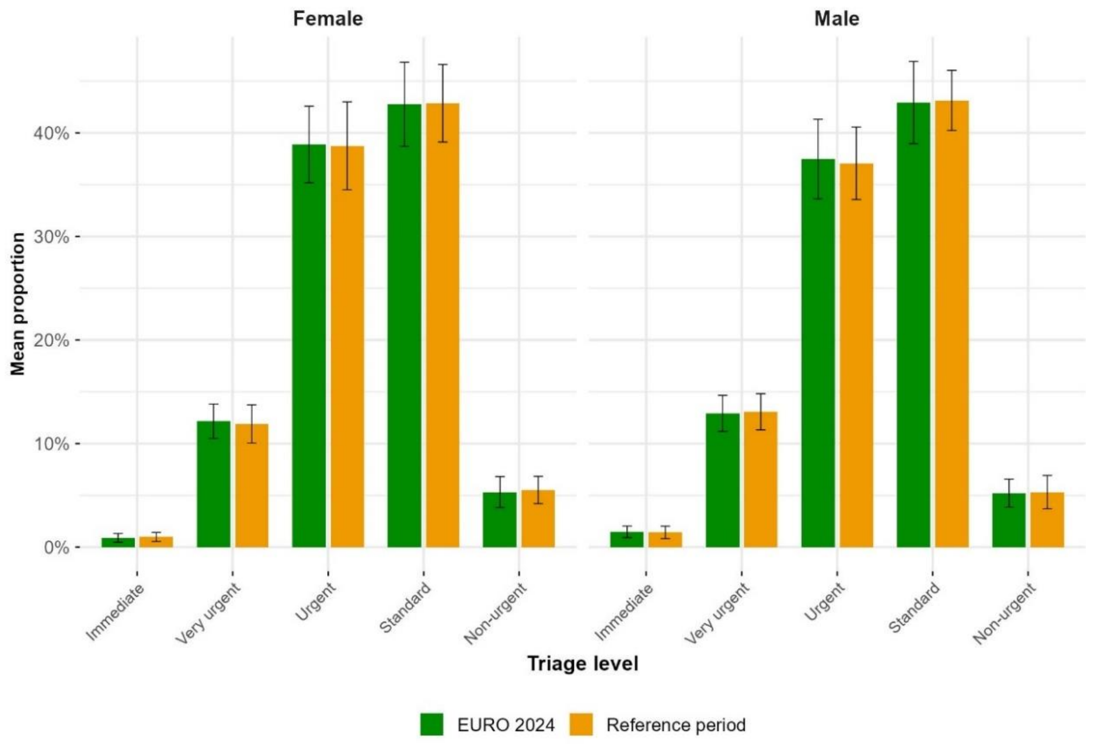

**Supplementary Figure S4:** Hourly attendance on the Germany game held on the 23rd of June, compared to a reference range representing two standard deviations above and below the mean of reference days. Game periods are highlighted.

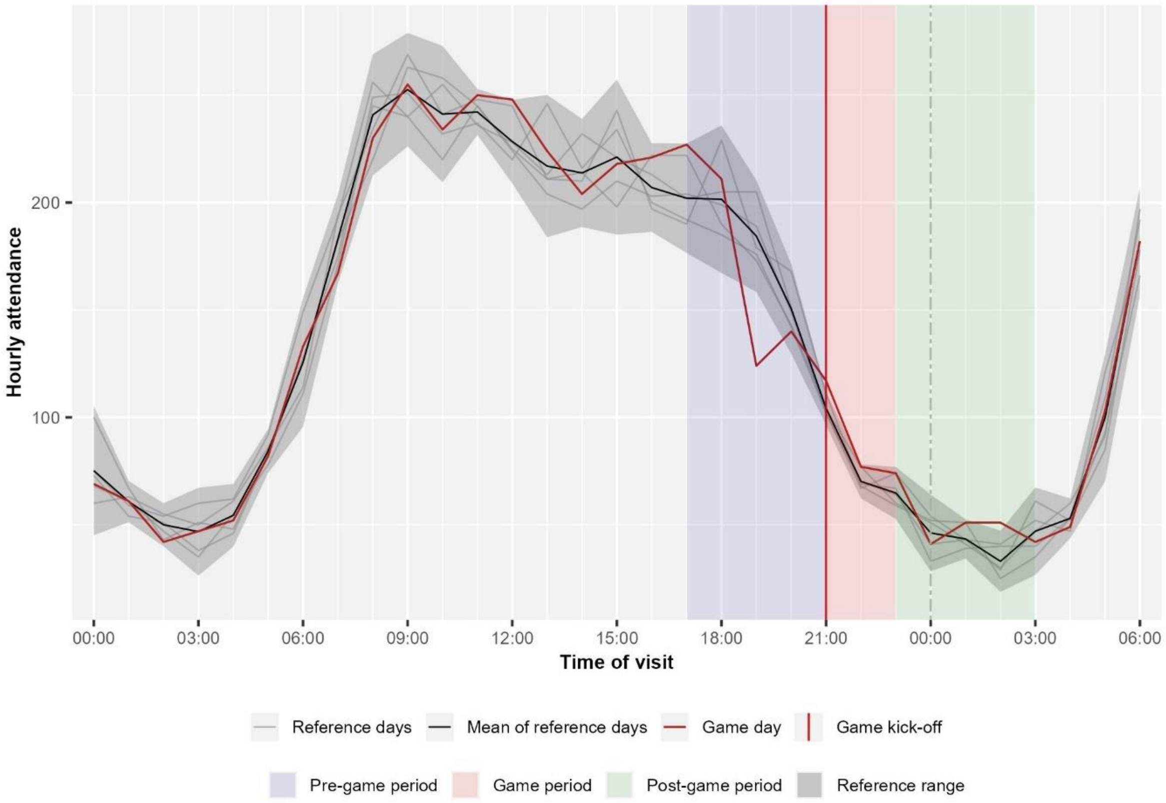


**Supplementary Figure S5:** *Hourly attendance on the Germany game held on 29th of June, compared to a reference range representing two standard deviations above and below the mean of reference days. Game periods are highlighted.*

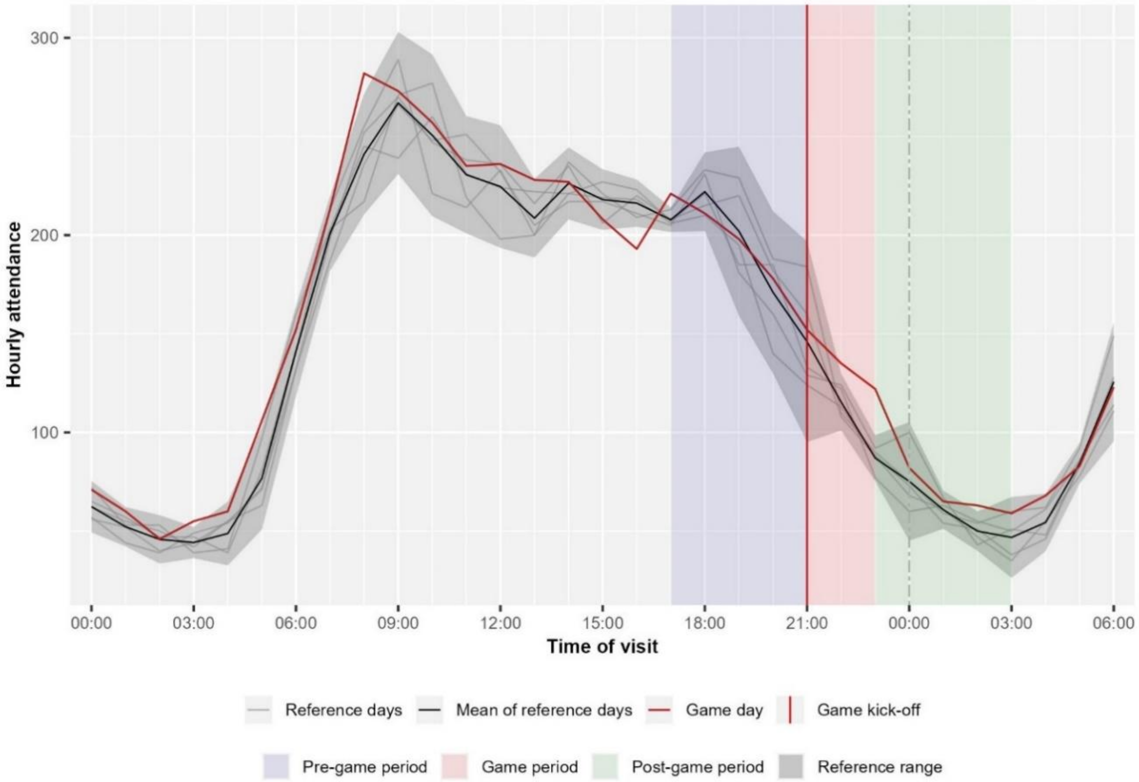

**Supplementary Figure S6:** *Hourly attendance on the Germany game held on 5th of July, stratified by age group, compared to a reference range representing two standard deviations above and below the mean of reference days. Game periods are highlighted.*

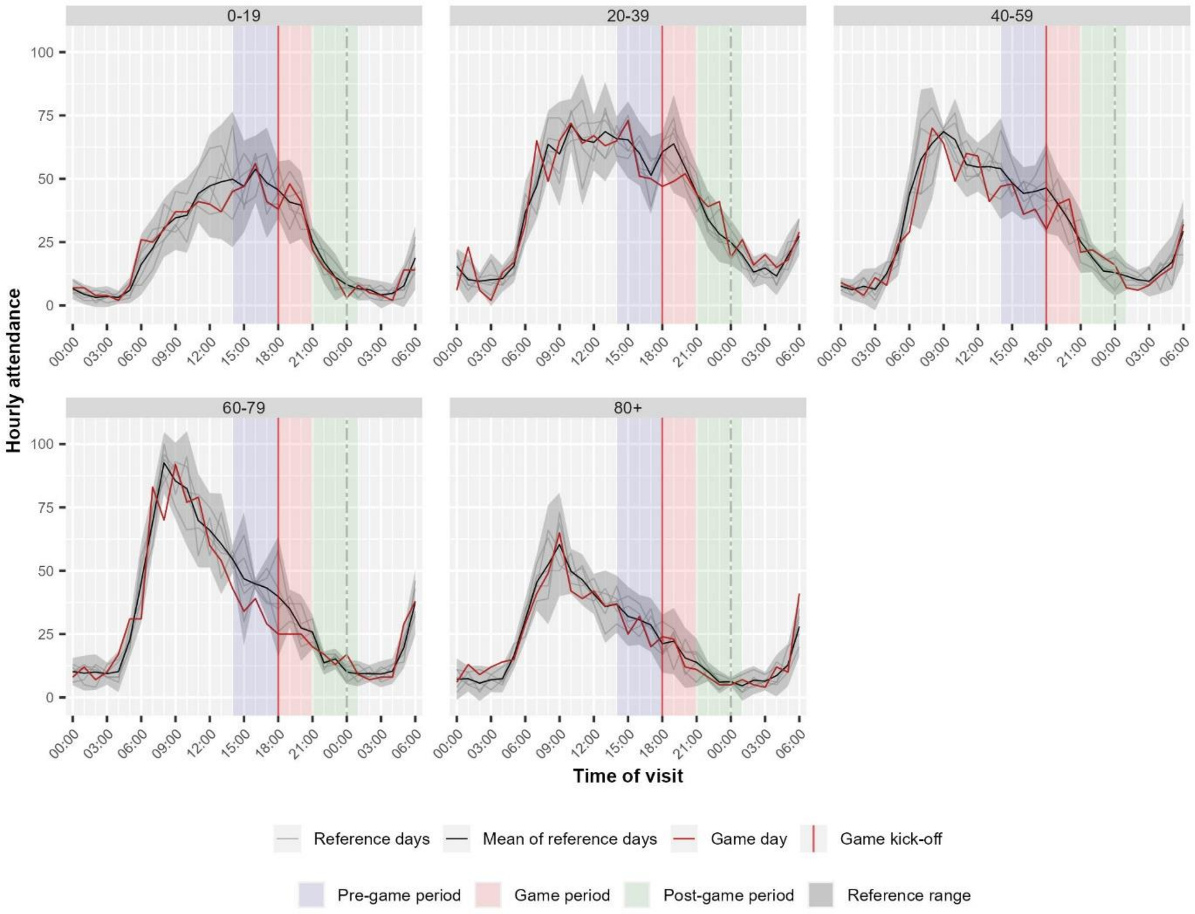

***Supplementary Figure S7:*** *Hourly attendance on the Germany game held on the 5th of July 2024 stratified by gender, compared to a reference range representing two standard deviations above and below the hourly mean of reference days. Game periods are highlighted*.

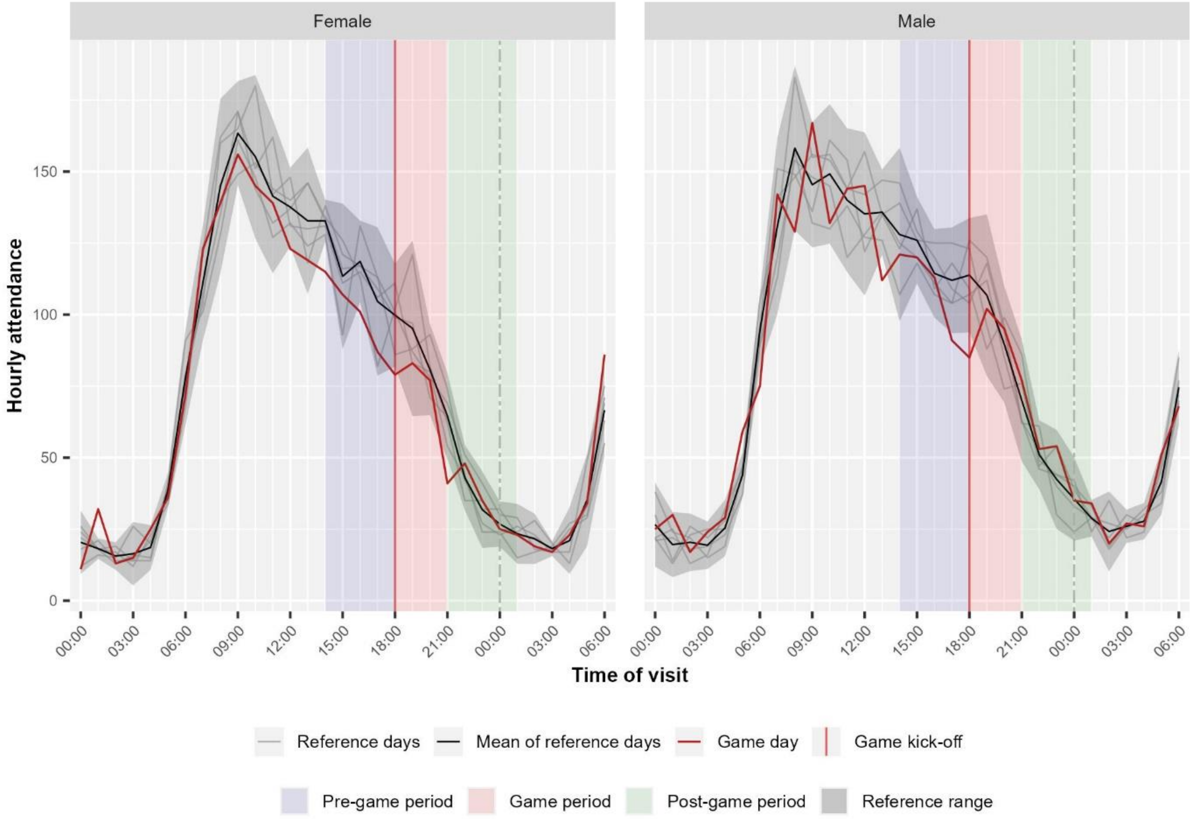
